# Genetics of Recurrent Miscarriage and Pregnancy Loss in Women’s Reproductive Health

**DOI:** 10.1101/2025.02.15.25321247

**Authors:** Zebinisa Mirakbarova, Vincent Pascat, Surayyo Akramkhanova, Chia-Yi Chu, Ulugbek Yusupov, Chiara Scapoli, Abdushukur Rakhmatullaev, Yuliya Kapralova, Sevara Nishanova, Mehribon Nazirova, Gulsanam Atamurotova, Konstantin Rudometkin, Maftuna Sodiqova, Lutfiya Karimova, Gulnoza Esonova, Hurshid Meylikov, Marguba Rejapova, Feruza Nishanova, Abrorjon Abdurakhimov, Inga Prokopenko, Dilbar Dalimova, Shahlo Turdikulova, Yevheniya Sharhorodska, Alisher Abdullaev

## Abstract

Adverse pregnancy outcomes, such as sporadic and recurrent miscarriages and stillbirths, are significant medical concerns, impacting up to 15% of clinically recognised pregnancies. These outcomes are highly complex and multifactorial, with up to 50% of cases classified as idiopathic, highlighting a substantial gap in our understanding of their biological basis. Along with external risk factors, polygenic variability contributes to idiopathic pregnancy loss, suggesting that large-scale genetic studies could offer insights into its mechanisms, reveal novel drug targets, and lead to new treatments. This study assesses current knowledge from genome-wide association studies (GWAS) using genotyping arrays, whole-genome imputation, and sequencing for variant discovery, emphasising genetic predisposition to adverse pregnancy outcomes. We summarise existing efforts identifying 30 genetic loci associated with pregnancy loss and related endophenotypes, integrating them into a polygenic score (PGS) and conducting a phenome-wide PGS association analysis of 280 ICD-10 outcomes in nearly 500,000 UK Biobank participants. We report associations between pregnancy loss PGS and an increased risk for diaphragmatic hernia (OR[95%CI]=1.02[1.01–1.03], P=9.15×10⁻⁷), eosinophilic esophagitis (OR[95%CI]=1.05[1.03–1.06], P=1.44×10⁻⁶), and asthma with exacerbation (OR[95%CI]=1.02[1.01–1.03], P=1.71×10⁻⁵), significant after correction for multiple testing and suggesting new mechanistic pathophysiology in pregnancy loss susceptibility. Additionally, Mendelian Randomisation (MR) studies identified higher BMI and smoking as risk factors for pregnancy loss, while the roles of caffeine and alcohol intake, maternal age, and family history of miscarriage warrant further investigation through adequately powered MR analyses. Well-designed and comprehensive GWAS studies, particularly across diverse ancestry groups, are urgently needed for idiopathic recurrent pregnancy loss. Such studies should overcome issues with identification of women suffering for this condition and related pregnancy losses to support better care and timely interventions, aiming for healthy live birth outcomes.

## Introduction

Pregnancy loss (PL) is a frequently infertility-related condition, affecting approximately 0.8% to 1.4% of women in the general population. PLs are subdivided into sporadic miscarriage (a single miscarriage before the 24th week of gestation), recurrent miscarriage (two or more consecutive unexplained miscarriages before the 24th week of gestation, varying by country definition and application), and stillbirth (loss of a foetus after the 24th week of gestation) (Dimitriadis et al., 2020; Rpl et al., 2018). PL management at country level necessitates comprehensive healthcare interventions and support to families and women. National health systems should allocate resources to provide timely and effective care, including mental health support and reproductive health services (Hennessy & O’Donoghue, 2024). Addressing PL is crucial for improving overall maternal health and well-being and reducing healthcare costs associated with complications and long-term care.

Miscarriage occurs in up to 15% of clinically detected pregnancies (Hong Li & Marren, 2018; Woolner & Bhattacharya, 2023). The majority, approximately 60%, are associated with sporadic chromosomal abnormalities, with trisomy being the most common and partially influenced by maternal age. These sporadic karyotypic abnormalities are most frequently found in products of conception, while the incidence of karyotypic abnormalities in parents is low. Balanced reciprocal translocations and Robertsonian translocations are reported for 2–5% of couples with recurrent PL (Practice Committee of the American Society for Reproductive, 2012).

While chromosomal abnormalities contribute significantly to PLs, other factors also play a critical role. Uterine structural anomalies are present in approximately 7–28% of women with recurrent PL (Carbonnel et al., 2021). Endocrine disorders, such as thyroid dysfunction and diabetes, are associated with 12-20% of miscarriages (Quenby et al., 2021). Severe infections and chronic endometritis are potential causes of PL, as the release of anti-inflammatory cytokines by placental and decidual tissues in response to these infections can disrupt pregnancy (Baqer et al., 2022; Pirtea et al., 2021). Antiphospholipid syndrome, an autoimmune condition, is reported in 8-42% of women with recurrent PL. Inherited thrombophilia’s, particularly factor V Leiden (FVL) mutations occur in 1-10% of cases. The prothrombin gene (PG) mutations contributing to RPL were found among 2–4% of European Caucasians. Overall, FVL and PG mutations may increase the risk of RPL by 2.44-fold and 2.08-fold, respectively. The prevalence of antiphospholipid antibodies is estimated at 15–20% among women with RPL (Regan et al., 2023). The Antiphospholipid syndrome triggers an inflammatory response due to antiphospholipid antibodies targeting the vascular endothelium and chorionic/placental cells, which promotes thrombosis (Dimitriadis et al., 2020; Liu et al., 2021; Sergi et al., 2015).

Recent European Society of Human Reproduction and Embryology (ESHRE) guidelines emphasised the importance of a **personalised prognosis**, which integrates a comprehensive evaluation of maternal age and full reproductive history, including the number, sequence, and outcomes of previous pregnancies. This individualised approach helps to inform management strategies and treatment options for women experiencing PL. However, for ***half of RPL patients, the causes remain unclear***, classifying them as idiopathic. Such adverse PL outcomes reflect a complex and multifactorial condition.

The role of maternal and paternal common genetic factors in idiopathic (recurrent) PL remains poorly understood. Prior to the release of the complete human genome, genetic studies reported effects of DNA variants in human major histocompatibility complex, at *CTLA4*, oestrogen receptor, and luteinizing hormone genes on PL (Christiansen et al., 1996; Taylor et al., 1993; Tsai et al., 1998; Tulppala et al., 1998). Although the role of genetic variants in pathophysiology of PL phenotypes remains to be described, emerging genetic/genomic technologies have started to bring novel insights into the role of DNA polymorphisms in PL pathogenesis. Despite huge advancement of knowledge for other complex conditions, such as obesity and diabetes, success from genome-wide association studies (GWAS) and next-generation sequencing studies, as well as the functional characterisation of associated loci for PL outcomes remains scarce. ***Slow advancement of knowledge about genetic underpinnings of PL is likely due to the highly heterogeneous nature of the condition, as well as challenges in data collection about women suffering from miscarriage(s) in both targeted studies and electronic health records*.**

In this study, we integrate current knowledge from large-scale GWAS, synthesising information about genetic loci implicated in ***idiopathic PL and its endophenotypes*** and evaluate suggested pathophysiological processes that may affect individual susceptibility.

### Miscarriage and stillbirth through the lens of GWAS

We focussed on large-scale genetic studies and GWAS dissecting susceptibility to recurrent miscarriages or stillbirth. The emergence of GWAS in 2007 (Klein et al., 2005; Manolio et al., 2009; Morales et al., 2017) allowed the identification of millions of variants across the human genome (Tam et al., 2019; Tosto & Reitz, 2013). The first GWASs on reproductive losses were published in 2019, only after a decade of GWAS successes for other phenotypes, with the majority of participants being of Western European ancestry, and minor representation from African and Asian populations (Fang et al., 2019). We queried miscarriage GWAS on the GWAS catalogue, PubMed, biorxiv/medrxiv, and Google Scholar from 2018 to Sept 2024, using a range of search item combinations: GWAS, sporadic miscarriage, recurrent miscarriage, spontaneous abortion, missed abortion, recurrent PL, self-reported PL, RPL, multiple consecutive miscarriage, stillbirth (**Table 1**).

**Table 1.** Published large-scale genetic studies for pregnancy loss outcomes.

| Phenotype | Sample size | Type of genetic data | Ancestry | Reference |
| --- | --- | --- | --- | --- |
| Sporadic miscarriage | 340,751 sporadic miscarriage cases, 570,138 controls | GWAS | Transethnic/<br>European | Reynoso et al., 2024 [66] |
| Recurrent miscarriage | 50,861 recurrent miscarriage cases nested within sporadic miscarriage cases, 570,138 controls | GWAS | Transethnic | Reynoso et al., 2024 [66] |
| Spontaneous abortion, missed abortion recurrent pregnancy loss, or self-reported pregnancy loss | 114,761 cases; 565,604 controls | GWAS | European | Steinthorsdottir et al., 2024 [65] |
| Gestation length, preeclampsia, gestational diabetes mellitus, pregnancy loss | 10,038 nulliparous cases | GWAS | European/<br>African/<br>Admixed<br>American | Khan et al., 2024 [63] |
| Sporadic miscarriage | 69,054 cases; 359,469 controls | GWAS | Transethnic | Laisk et al., 2020 [24] |
| Multiple consecutive miscarriage | 750 cases 359,469 controls | GWAS | European | Laisk et al., 2020 [24] |
| Stillbirth | 1,008 Tibetan women | WGS | Asian | Jeong et al., 2018 [57] |

One of the biggest miscarriage GWAS to date involved 69,054 cases from five ancestries for sporadic miscarriage and 750 cases of European ancestry for multiple consecutive miscarriages (five and above consecutive miscarriages), along with up to 359,469 female controls (Laisk et al., 2020). This study identified variants near *MIR148A* and *DDX39AP1* associated with sporadic miscarriages in the Transethnic level and European population respectively, while variants in *NAV2*, *LINC01679*, *NBAS*, and *RPS20P25* were associated with multiple consecutive miscarriages. Notably, all identified variants except one were rare (MAF < 1%), and only one variant was of low frequency (2% < MAF < 5%), which is uncommon for large-scale GWAS efforts that generally rely on the common phenotype-common variant hypothesis for statistical analysis (Claussnitzer et al., 2020).

Laisk et al. study highlighted the potential role of microRNAs (miRNAs) in susceptibility to miscarriage (Laisk et al., 2020). miRNAs are messengers from DNA to protein, they are invaluable in regulating gene expression by binding to complementary sequences in messenger RNA (mRNA) molecules and help defining cell types and guiding their development. Little is known to date about the role of *MIR148A* and *LINC01679* in reproductive processes, but recent studies suggest that high levels of *MIR148A* disrupt B cell tolerance by helping immature B cells survive by lowering the levels of proteins that normally promote cell death (tumour suppressors) and prevent autoimmunity (Gonzalez-Martin et al., 2016). The role of DEAD-Box Helicase 39A Pseudogene 1 (*DDX39AP1*) in pregnancy aetiology is also unclear, although previous GWAS associated *DDX39AP1* with atrial fibrillation and stroke, pathological myopia and cholesterol change in response to fenofibrate in statin-treated T2D (Fan et al., 2014; Rotroff et al., 2018). MSC-associated *NAV2* protein is essential for neurite outgrowth and axonal elongation, acting as a key regulator of cranial nerve outgrowth and blood pressure (McNeill et al., 2010; Muley et al., 2008). It was also shown, that, through activating the Wnt/β-catenin signalling pathway, NAV2 promotes the proliferation and invasion of fibroblast-like synoviocytes (FLSs) in rheumatoid arthritis (RA) and colorectal cancer (Tan et al., 2015; Wang et al., 2021).

The study by Laisk et al. identified a genome-wide significant association with the *TLE1* gene (rs146350366) for sporadic miscarriage in their European ancestry meta-analysis (Laisk et al., 2020). TLE1 protein was also reported to inhibit Wnt signalling, other cell fate determination signals and the differentiation of neural progenitor cells also acting as a corepressor for NF-kB, regulating inflammation and potentially influencing various diseases, including cancers (sarcoma, mesothelioma), yet the exact mechanisms remain unclear (Dastidar et al., 2012; Terry et al., 2007; Tetsuka et al., 2000; Yuan et al., 2016). *TLE1* is crucial for the survival of postmitotic neurons, functioning through interactions with *FoxG1* and pathways like CK2 and *PI3K*-*Akt* (Yuan et al., 2016). *NBAS* gene encoded Neuroblastoma amplified sequence protein is characterised by two leucine zipper domains, a ribosomal protein S14 signature domain, and a Sec39-like domain (Ben Hadj Hmida et al., 2016). NBAS is essential for maintaining Golgi-ER transport integrity and is critical for normal immune cell function. Mutations in the *NBAS* gene have been associated with various disorders, such as infantile liver failure syndrome type 2 (ILFS type 2) and hemophagocytic lymphohistiocytosis (HLH) (Bi et al., 2022; Chavany et al., 2020; Hasosah et al., 2017; Ji et al., 2023).

Using a relatively small sample size, another polygenic score-based study evaluated the predictive ability of a composite DNA variant set implicated in various biological pathways, identifying a variant within the *IL10* gene associated with RPL (OR 1.60 (95% CI: 1.07-2.42)) (Loizidou et al., 2021). Furthermore, several small-scale case-control studies associated variants in *IL10* gene with recurrent PL and spontaneous abortions (L. Bohiltea & E. Radoi, 2014; Qaddourah et al., 2014; Su et al., 2016; Sudhir et al., 2023). Experimental studies on rats showed that interleukin-10 (IL-*10*) significantly reduces the foetal death rate and growth restriction induced by low-dose endotoxin exposure. This effect is possibly due to the suppression of TNF-alpha and nitric oxide-mediated cell death in the uteroplacental unit (Rivera et al., 1998). Validation of these findings in large-scale GWAS for PL would still be required.

For many years the intronic regions were considered as an “exon keeping and separating blocks”, while intronic variants were undervalued. Studies in the last decades show their functional activity in mRNA processing (Chorev & Carmel, 2012), splicing, and regulation of expression (Choi & Chan, 2015). Furthermore, introns may serve as targets for mutations at a significantly higher proportion of mutational hotspots due to their arrays of essential functional elements, including intron splice enhancers and silencers, trans-splicing elements, and other regulatory components (Gingeras, 2009; Solis et al., 2008; Tress et al., 2007; Wang et al., 2009). Although GWA methods made it easier to identify intronic variants and computational tools are emerging to assess their potential impact on disease and phenotypes, understanding their role in complex traits remains a challenge due to the limitations of traditional variant interpretation methods in analysing functional effects of intronic variants (Barbosa et al., 2023; Chorev et al., 2017; Lin et al., 2019).

Although GWAS started over 15 years ago, the number of genome association studies focusing on individuals of Asian descent is limited. To date, three relatively small GWAS have been conducted for the adverse pregnancy outcomes for women of Asian ancestry. The first GWAS focused on adaptive evolution markers in 1,008 Tibetan women aged 39 and over, considering they had completed reproductive potential. The study identified 14 associated variants between *VRK1* and *PAPOLA* gene (**Table 2**), encoding a poly-A tail polymerase (Uniprot ID: P51003), a key component of RNA processing, expressed in the nucleoplasm (Jeong et al., 2018). PAPOLA protein is a key input for the intronless pre-RNA cleavage complex. A study on a limited number of patients in Japan, associated *MEIG1* with miscarriage (**Table 1**) along with five other potential SNVs (Yoshihara et al., 2022). Several functional studies previously reported the role of *MEIG1* alterations in male infertility in mice models and humans due to impaired spermatogenesis (Li et al., 2015; Li et al., 2021; Zhang et al., 2018; Zhang et al., 2017).

**Table 2.** Genome-wide studies and genetic loci associated with pregnancy loss outcomes.

| Chromosome: position | Nearby gene | RSID | Risk allele/ Other allele | Risk allele frequency | Risk allele effect estimate: OR/RR (95% CI)* or beta^, where reported | Phenotype: SM - sporadic miscarriage; RM - Recurrent miscarriage; MCM - Multiple consecutive miscarriage; GDM - Gestational diabetes mellitus | Reference |
| --- | --- | --- | --- | --- | --- | --- | --- |
| 1:50494849 | *FAF1* | rs10888690 | C/T | 0.430 | 1.021 (1.015 – 1.029)‡ | SM | Reynoso et al., 2024 |
| 1:150491055 | *TARS2* | rs66499095 | D/I | 0.196 | 1.024(1.016 – 1.031)‡ | SM | Reynoso et al., 2024 |
| 2:15017885 | *NBAS* | rs138993181 | A/C | 0.006 | 3.61 (2.23-5.84)* | MCM | Laik et al., 2020 |
| 2:60426705 | *BCL11A* | rs11125830 | A/G | 0.014 | 1.043 (1.027 -1.057) ‡ | RM | Reynoso et al., 2024 |
| 2:139844192 | *LRP1B* | rs13421417 | A/T | 0.311 | 1.019(1.013 – 1.026)-‡ | SM | Reynoso et al., 2024 |
| 3:114815630 | *ZBTB20* | rs72956265 | A/G | 0.02 | 3.26(2.15 - 4.95) ‡ | GDM | Khan et al., 2024 |
| 3:117597695 | *LINC03051* | rs9875219 | T/G | NA | 1.043(1.028 - 1.059) ‡* | RM | Reynoso et al., 2024 |
| 4:102267552 | *SLC39A8* | rs13107325 | T/C | 0.758 | 1.040 (1.028 - 1.053) ‡ | SM | Reynoso et al., 2024 |
| 4:183047617 | *TENM3* | rs756996932 | NA | NA | 2.5 (1.858 - 3.364) ‡ | RM | Reynoso et al., 2024 |
| 5:166060445 | *RPL7P20* | rs79596863 | G/A | 0.04 | 2.75 (1.95 – 3.89) ‡ | GDM | Khan et al., 2024 |
| 7:25799726 | *MIR148A* | rs10270417 | C/T | 0.017 | 1.0(0.9–1.0)* | SM | Laisk et al., 2020 |
| 8:8401607 | *PRAG1* | rs2920991 | A/T | 0.501 | 1.018 (1.012 - 1.025) ‡ | SM | Reynoso et al., 2024 |
| 8:9418515 | *TNKS* | rs35847492 | C/G | 0.293 | 1.021(1.014- 1.029) ‡ | SM | Reynoso et al., 2024 |
| 8:10739038 | *PINX1* | rs12334416 | A/C | 0.468 | 1.018(1.012 – 1.025) ‡ | SM | Reynoso et al., 2024 |
| 9:81063077 | *TLE1* | rs7859844 | A/G | 0.064 | 1.68 (1.42-1.99)* | MCM | Laisk et al., 2020 |
| 11:19610850 | *NAV2* | rs143445068 | C/G | 0.008 | 3.43 (2.35-5.00)* | MCM | Laisk et al., 2020 |
| 11:106675610 | *GUCY1A2* | rs10890563 | C/T | 0.11 | 1.94 (1.52 - 2.45) ‡ | GDM | Khan et al., 2024 |
| 13:22779354 | *DDX39AP1* | rs146350366 | G/A | 0.012 | 1.4 (1.2–1.6)* | SM | Laisk et al., 2020 |
| 13:58814438 | *DIAPH3* | rs1855263 | G/A | 0.672 | 1.020 (1.013 - 1.027) ‡ | SM | Reynoso et al., 2024 |
| 14:97131992 | *PAPOLA* | rs1957819 | T/C | 0.083 | NA | Stillbirth | Jeong et al., 2018 |
| 15:93045079 | *RGMA* | rs62021480 | G/C | 0.08 | 3.53 (2.43 – 5.14) ‡ | Pregnancy loss | Khan et al., 2024 |
| 16:9212279 | *C16orf72* | rs10852372 | C/A | 0.592 | 1.019 (1.012 - 1.026) ‡ | SM | Reynoso et al., 2024 |
| 16:84329456 | *WFDC1* | rs2550487 | A/C | 0.01 | -1.28 ‡^ | Gestation length | Khan et al., 2024 |
| 19:13015347 | *SYCE2* | rs189296436 | A/NA | 0.18–1.27 | 1.22 ( 1.16–1.30) | Spontaneous abortion, missed abortion recurrent pregnancy loss, or self-reported pregnancy loss | Steinthorsdottir et al., 2024 |
| 19:31414662 | *TSHZ3* | rs2032905 | G/A | 0.577 | 1.020 (1.013 - 1.026) ‡ | SM | Reynoso et al., 2024 |
| 21:43412110 | *LINC01679* | rs183453668 | A/G | 0.005 | 3.78 (2.44-5.85)* | MCM | Laisk et al., 2020 |
| 22:46351905 | *TRMU* | rs142795512 | T/C | 0.02 | 5.31 (3.11 – 9.06) ‡ | Pregnancy loss | Khan et al., 2024 |
| X:119710269 | *AC005052.1* | rs58548906 | G/A | 0.0099 | -1.43^ | Gestation length | Khan et al., 2024 |

‡ - threshold for significance ( $P\text{-value} < 10^{-8}$ ) more stringent compared to other studies with classical multiple testing correction for genome-wide significance $p\text{-value} < 5 \times 10^{-8}$
\*- analyses in European ancestry populations
^ - effect size measure is beta.

Recently, three larger GWAS for miscarriage were published, focusing primarily in population of European ancestry and transethnic populations. One of these studies, published in 2024, focused on association study of PL, gestational diabetes and gestational length in a cohort of 10,038 nulliparous women of European and African descent, and reported seven variants, reaching genome-wide significance. The single nucleotide polymorphism (SNP) rs62021480 reported with minor allele frequency of 0.048-0.18 leads to synonymous mutation in the *RGMA* gene, which potentially influences transcription factor binding, exerts post-transcriptional regulatory effects is linked to an increased likelihood of experiencing poorer kidney function or damage as evidenced in kidney disease study (p=0.26) (Khan et al., 2024; Mota-Zamorano et al., 2021). The functional and structural effects of an intronic variant, rs142795512, with the highest OR for PL outcome (OR=5.31) have not been previously identified and are based on bioinformatics findings and previous studies of other variants in the *TRMU* gene and its expression levels (Khan et al., 2024). This variant could regulate the expression levels of the *TRMU* gene in complex with other variants, but evidence support for these claims is limited and no experimental data is yet available on downstream biological mechanisms.

A study of 114,761 cases and 565,604 controls from the Northwestern Europe (The United Kingdom, Denmark, Finland, Iceland) and the USA population revealed association of the single missense variant rs189296436 with PL. rs189296436 leads to His89Tyr mutation in SYCE2 protein (Steinthorsdottir et al., 2024). Although the specific role of this variant has not been fully explored, studies suggest that *SYCE2*:p.His89Tyr variant, interferes with efficient synapsis and recombination during meiosis by disrupting the structural assembly of the SYCE2–TEX12 synaptonemal complex, as this variant is located at a critical protein-protein interaction site essential for the assembly of the central element of the synaptonemal complex.

The most recent and largest GWAS identified **nine independent loci** associated with sporadic miscarriages in European and transethnic studies, two variants linked to recurrent miscarriage in the European population, and one variant associated with recurrent miscarriage in the transethnic cohort (Reynoso et al., 2024). Variants rs9875219, rs756996932, and rs11125830 in *LINC03051*, *TENM3* and *BCL11A* loci respectively, associated with recurrent miscarriages, located in intronic and intergenic non-coding regions, have not yet been investigated in subsequent studies. Despite the association of nearby genes (*TENM3*, *LINC03051*) with bone development and syndromic microphthalmia, the role of these variants in pregnancy pathogenesis is undetermined (Pei et al., 2016; Zhou et al., 2022). Notably, rs11125830 was previously in linkage disequilibrium with other variants associated with atrial fibrillation and stroke (Holm & Gudbjartsson, 2012).

### PL polygenic score and health outcomes in the UK Biobank

We conducted a PheWAS to systematically evaluate associations between a miscarriage polygenic score (PGS) and various related health outcomes using data from the UK Biobank. Using information about 30 DNA variants from PL GWASs as described above (**Table 2**), we built an unweighted PGS for PL in the UK Biobank, including 459,009 individuals of multiple ancestry. Briefly, each independent SNV (LD r²<0.2) was assigned a weight of 1 if the effect allele was associated with heightened risk of miscarriage. The UK Biobank imputed genotype dataset was filtered standard quality control measures (filtering individuals and SNs with an information score greater than 0.4). We extracted all available the International Classification of Diseases-10 (ICD-10) code disease outcomes from the UK Biobank hospital records up to December 2023. We performed a phenome-wide association analysis (PheWAS) between the PL PGS and 280 ICD-10 phenotypes (***Supplementary Table 1***) using *comorbidPGS* v.0.3 *R-studio package (Pascat et al., 2024)*. The resulting associations were corrected for multiple testing using Bonferroni correction (P-valueBonferroni=0.05/280=1.79×10^-4^).

We identified three significant associations with the 30 SNP PL PGS. Most significant association was with higher risk of **diaphragmatic hernia** (OR [95%CI]=1.02 [1.01 – 1.03], P-value=9.15×10^-7^), ICD-10 code K449, observed in 50,001 individuals of UK Biobank. Second most significant association was with **eosinophilic esophagitis (EoE)** (OR [95%CI] = 1.05 [1.03–1.06], P-value=1.44×10^-6^), ICCD-10 code K20, reported for 12,275 UK Biobank individuals. The third most significant PheWAS association was for **asthma with exacerbation** (OR[95%CI]=1.02[1.01–1.03], P-value=1.71×10^-5^), ICD-10 code J459, recorded in 41,495 cases, a respiratory condition (**Figure 1**). Additionally, we observed trends of association (1.79×10^-4^<P-value<1.79×10^-3^) with hypothyroidism (indicating an inverse relationship, where a higher miscarriage PGS was associated with decreased risk of hypothyroidism), urinary retention (associated with drug use), malignant neoplasms of the genital organs, and allergy status to penicillin. The three significantly associated with PL PGS conditions in UK biobank suggest potential for new pathophysiology of this idiopathic women health outcome.

**Figure 1.**
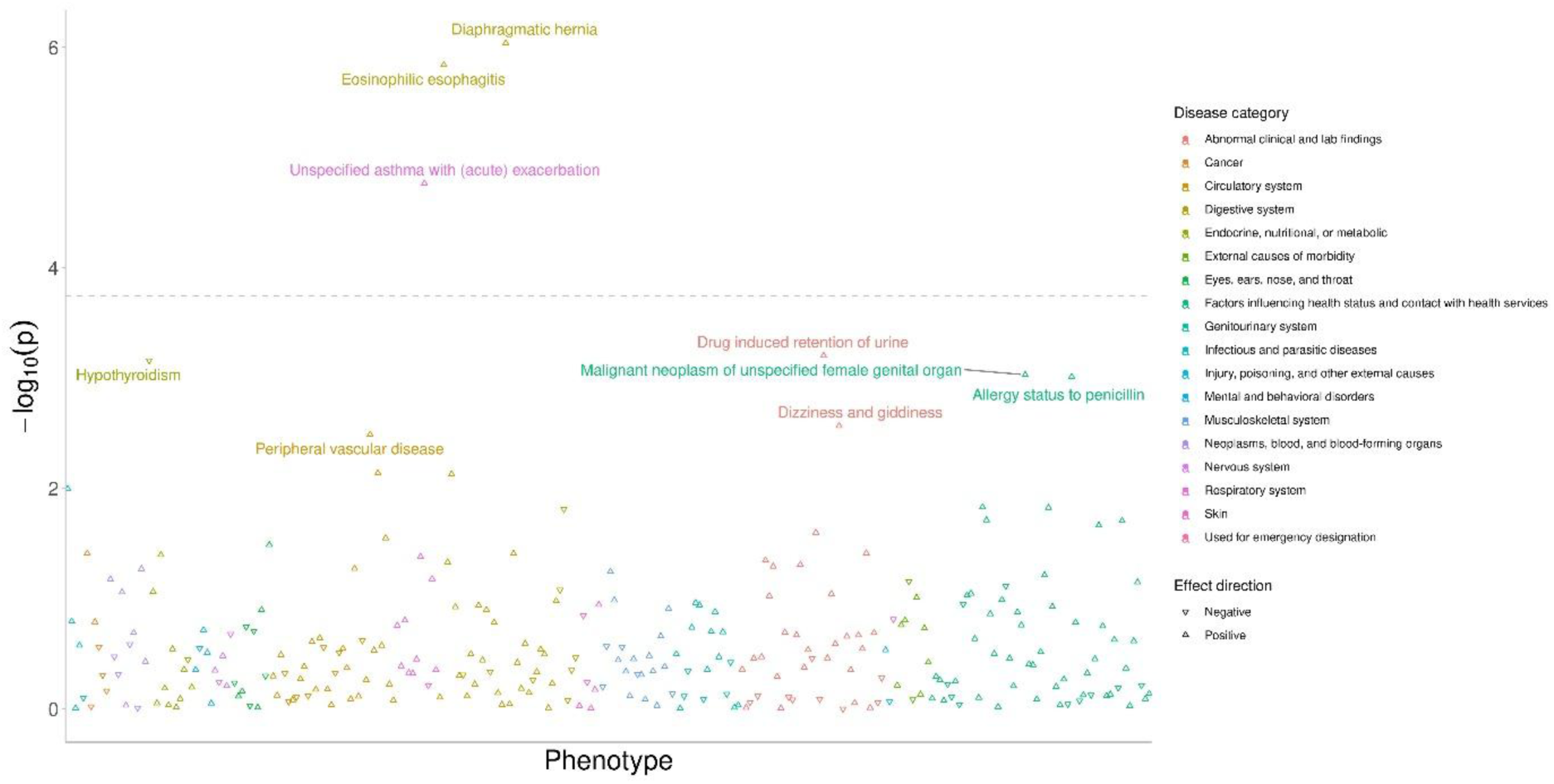
PheWAS between pregnancy loss PGS and 280 ICD-10 defined health outcomes in UK Biobank

**Diaphragmatic hernia** (DH) is a herniation of abdominal structures within thoracic cavity through a defect on the diaphragm. Diaphragmatic hernia is typically a congenital condition, but there is a growing number of acquired case instances, which have non-congenital origin. The reported incidence of diaphragmatic hernia is approximately 0.8 to 5 out of 10,000 live births (Chandrasekharan et al.). Acquired diaphragmatic hernia can occur due to thoracoabdominal trauma, surgical complications, or other factors occurs in approximately 0.8 to 3.6% of cases, with the incidence of herniation following this being relatively low. In UK Biobank we identified a much higher rate in adult life, which is related to an increasingly common problem in Westernised developed countries, a gastroesophageal reflux disease (GERD). Acquired diaphragmatic hernia and PL could share a complex pathogenesis that involves several interconnected factors. When an acquired diaphragmatic hernia occurs, often due to trauma, it can lead to significant physiological stress which triggers systemic inflammation and can negatively impact placental function, disrupting the delicate balance necessary for healthy foetal development. Respiratory distress caused by a hernia can further complicate pregnancy by reducing the oxygen supply to the foetus. This reduction can increase the risk of foetal hypoxia, which is associated with PL or other adverse outcomes. On the other hand, **congenital diaphragmatic hernia** (CDH) is a birth defect (up to 1/2,000 life birth in the UK and potentially more frequent in idiopathic PL) characterised by the incomplete closure of the diaphragm, allowing foetal abdominal organs to herniate into the chest cavity (Long et al., 2018; Long et al., 2019). This condition can lead to high rates of intrauterine mortality. The exact cause of most cases of CDH remains unclear, but it is increasingly evident that genetic factors play an important role in many CDH instances.

CDH can present as an isolated birth defect or in association with other non–hernia-related anomalies. A substantial number of individuals with nonisolated CDH have chromosomal anomalies, which are also one of the leading causes of PL. Some cases of nonisolated CDH are linked to aneuploidy, where entire chromosomes are either deleted or duplicated, with common forms including trisomy 13, 18, 21, and occasionally tetrasomy 12p or Pallister–Killian syndrome (Pober, 2007). Structural chromosome anomalies, such as deletions, duplications, inversions, and translocations, are also frequently reported in individuals with nonisolated CDH, highlighting the impact of chromosomal abnormalities on both congenital disorders and adverse pregnancy outcomes (Scott, 2007).

Genetic syndromes are identified in approximately 10% of CDH cases, with Fryns syndrome (MIM 229850) being the most prevalent. Fryns syndrome is an autosomal recessive disorder characterized by a combination of congenital abnormalities, including anophthalmia (absence of one or both eyes), facial cleft, micrognathia (undersized jaw), ventriculomegaly (enlargement of the brain’s ventricles), and diaphragmatic hernia. Patients with Fryns syndrome often exhibit additional features such as limb anomalies, cardiac defects, and renal malformations. However, reports of individuals with Fryns-like phenotypes linked to chromosomal anomalies—such as duplication of 1q24-q31.2, deletion of the terminal region of 6q, 8p23.1, and 15q26, as well as partial trisomy 22—indicate that some cases of CDH attributed to this autosomal recessive syndrome may actually represent genocopies of the disorder(Clark & Fenner-Gonzales; de Jong et al.; Dean et al.; Krassikoff N Fau - Sekhon & Sekhon; Slavotinek A Fau - Lee et al.). Many of the syndromes associated with CDH exhibit specific Mendelian inheritance patterns, and in some instances, the location and identity of the causative gene(s) have been identified. Notable examples include *CDKN1C* (MIM 600856), *NSD1* (MIM 606681), *CHD7* (MIM 608892), *NIPBL* (MIM 608667), *SMC1A* (MIM 300040), *EFNB1* (MIM 300035), *WT1* (MIM 607102), *GPC3* (MIM 300037) and *CXORF5* (MIM 300170). Among mendelian forms of Fryns-like CDH, *WT1* gene is near the enhancer-located rs5030123 *WT1* gene locus, associated with multiple types of hernias, including non-syndromal DH, suggesting clear biological link (Campbell et al., 2022). The existence of genetic syndromes associated with CDH provides one of the strongest lines of evidence that genetic factors play a role in the development of this condition. Similarly, many genetic syndromes, linked to PL, also underscore the impact of CDH on foetal viability and pregnancy outcomes. The additional assumption that most CDH cases arise from a complex inheritance pattern, where a combination of genetic and environmental factors influences the final phenotype, aligns with the sporadic nature of the condition and the limited number of familial cases reported in the literature. Furthermore, environmental stressors—such as toxins or nutritional factors like vitamin A—may contribute to variations by impacting genetically susceptible individuals. The interplay between multiple genes and environmental influences may also help explain the association of CDH with certain chromosomal abnormalities (Holder et al., 2007).

The second most genetically PL PGS-associated phenotype in UK Biobank was **eosinophilic esophagitis (EoE)**, whose exact pathogenesis remains unclear but is thought to arise from an immune/allergic reaction to food or environmental antigens (such as dust mites, animal dander, pollen, and molds). Genetic predisposition, particularly oesophageal-specific variations, likely contribute to EoE’s complex heritability, as evidenced by familial patterns. Individuals with allergic conditions like atopic dermatitis, asthma, or other food and environmental allergies are at higher risk of EoE. The current understanding of EoE’s pathophysiology suggests that disrupted oesophageal epithelial barriers enable antigen exposure, triggering an upregulation of atopy-related cytokines. This leads to inflammatory cell infiltration and the activation of the eosinophilic esophagitis transcriptome, culminating in an inflammatory cascade. Over time, this cascade causes transmural injury, fibrosis, and narrowing of the oesophagus. Many genes implicated in EoE overlap with those involved in type 2 immune responses, such as in **asthma**, atopic dermatitis, and allergic rhinitis (Figure 1) (Gupta & Grinman). Notably, corticosteroids are effective in treating both EoE and asthma, supporting the hypothesis of a shared immunological mechanism rooted in inflammation. **Asthma**, the third most significantly associated with PL PGS in UK Biobank, similarly to EoE, involves persistent airway inflammation, with triggers like infections, allergens, and irritants that lead to multicellular inflammation, bronchial hyper-responsiveness, and airflow obstruction. Exacerbations of asthma are often characterized by the infiltration and activation of eosinophils, neutrophils, and lymphocytes, highlighting both inflammatory and immunological pathways. In cases of PL, the immune system’s balance is critical. Dysregulation, involving immune cell types (both innate and adaptive, such as NK cells, neutrophils, T cells, and B cells) (Wieseler-Frank et al., 2005) activated in both EoE and asthma, can lead to immune attacks on the embryo or placenta, compromising pregnancy success (Guan et al., 2024).

### Causal relationships between potential risk factors and miscarriage/stillbirth

The genetic underpinnings of adverse pregnancy outcomes including miscarriages and stillbirth and their associations with various traits show a multifaceted landscape of maternal health exposome (**Figure 2**). Investigations into causal genetic associations shed light on the potential contributors to miscarriage and stillbirth risk, encompassing factors ranging from lifestyle choices to inherent genetic predispositions.

**Figure 2.**
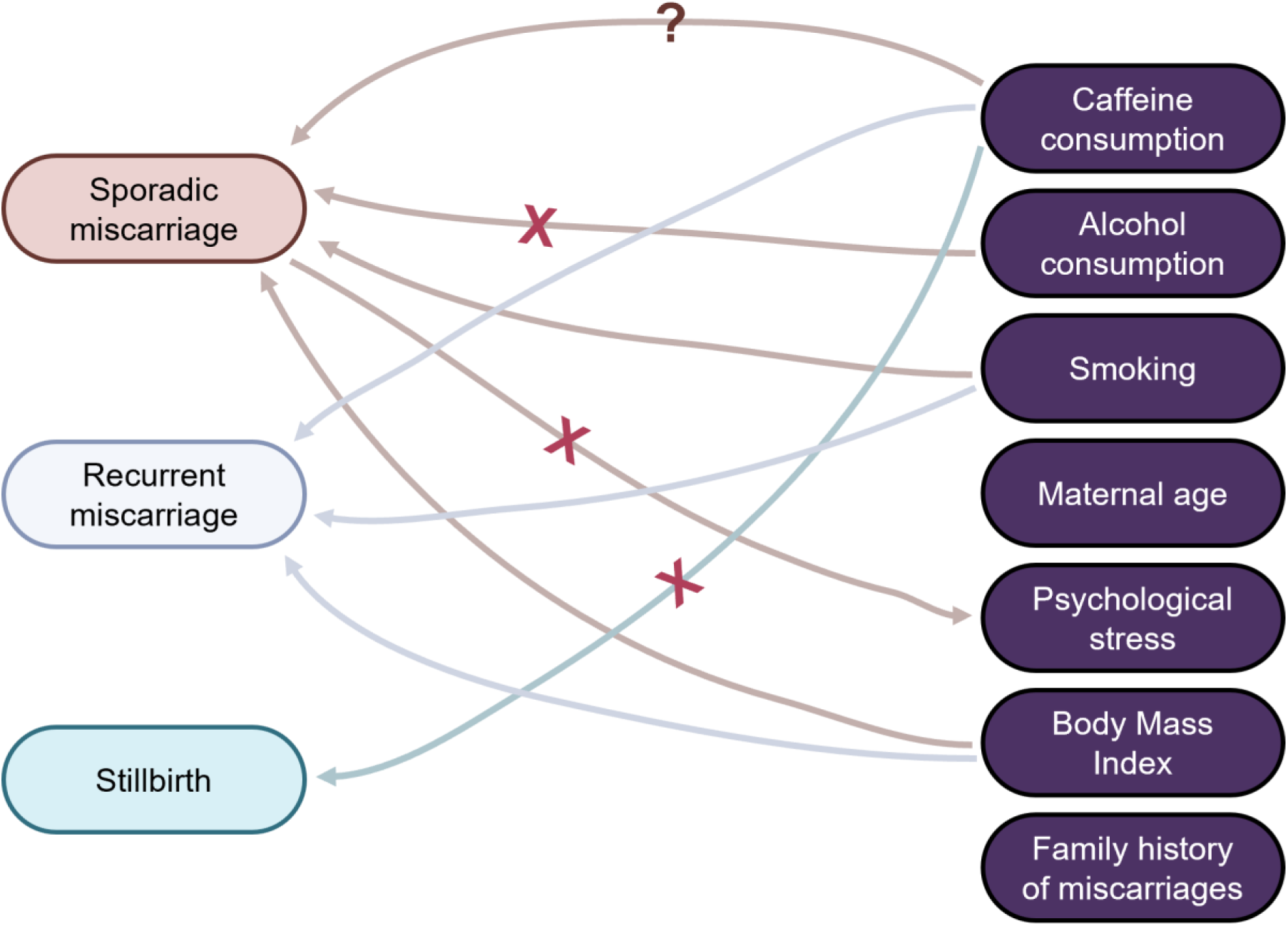
Schematic overview of causal relationships between adverse pregnancy outcomes and potential risk factors

**Smoking.** Smoking during pregnancy is a well-documented risk factor for adverse outcomes, including spontaneous miscarriage. Observational studies consistently show that smoking increases miscarriage risk, with a dose-response relationship (George et al., 2006; Pineles et al., 2014; Skogsdal et al., 2022). However, the association between smoking and recurrent miscarriage is less clear, with some studies indicating no significant link (Ng et al., 2021; Zhang et al., 2010). MR studies, however, provide strong evidence for a **causal relationship** between smoking and both sporadic and recurrent miscarriages, and stillbirth under the definition of PL, supporting the hypothesis that **smoking significantly increases miscarriage risk** (**Table 3**) (Laisk et al., 2020; Painter et al., 2021; Reynoso et al., 2024; Yuan et al., 2021).

**Table 3.** Mendelian randomisation analyses to dissect causal relationships between potential risk factors and pregnancy loss outcomes.

| Risk factor studied (exposure) | Pregnancy loss outcome | Number of SNPs used | Odds Ratio, OR (Confidence interval, CI) | P-value | Study size for outcome phenotype | Ancestry distribution for outcome phenotype | Reference |
| --- | --- | --- | --- | --- | --- | --- | --- |
| Caffeine consumption | Miscarriage recurrent | 127 SNPs | 1.48 (1.07, 2.04) | **0.019** | 52,087 cases/ 542,880 controls | 85% European, 11% Latinx, 4% African/ American | Reynoso et al., 2024 |
| Caffeine consumption | Miscarriage sporadic | rs1481012<br>rs4410790<br>rs9902453<br>rs7800944<br>rs1768<br>rs2472297<br>rs1260326<br>rs6265 | -0.048 (-0.138, 0.043) | 0.296 | 49,996 cases/ 174,109 controls | European | Nunes et al., 2023 |
| Caffeine consumption | Miscarriage sporadic | 127 SNPs | 1.27 (1.06, 1.52) | **0.013** | 334,593 cases/ 505,008 controls | 85% European, 11% Latinx, 4% African American | Reynoso et al., 2024 |
| Caffeine consumption | Stillbirth | rs1481012<br>rs4410790<br>rs9902453<br>rs7800944<br>rs1768<br>rs2472297<br>rs1260326<br>rs6265 | 0.006 (-0.012, 0.024) | 0.459 | 60,453 | European | Nunes et al., 2023 |
| Alcohol consumption | Miscarriage sporadic | 96 SNPs | 1.04 (0.89, 1.21) | 0.645 | 49,996 cases/<br>174,109 controls | European | Laisk et al., 2020 |
| Body Mass Index | Miscarriage sporadic | 265 SNPs | 1.06 (1.01, 1.12) | **0.024** | 50,060 cases/<br>174,109 controls | European | Venkatesh et al., 2022 |
| Depression | Miscarriage sporadic | 20 SNPs | 1.025 (0.915, 1.147) | 0.674 | 16,906 cases/<br>149,622 controls | European | Ling et al., 2024 |

**Adiposity (Body Mass Index, BMI).** Numerous epidemiological studies highlight a positive association between high BMI and miscarriage risk. Meta-analyses and case-control studies confirm that high BMI significantly increases the likelihood of miscarriage, potentially due to metabolic dysfunction and hormonal imbalances (Cavalcante et al., 2019; Jung et al., 2015). MR studies using genetic markers for BMI reinforce these findings, demonstrating a causal relationship between higher BMI and increased miscarriage risk (**Table 3**) (Venkatesh et al., 2022).

**Caffeine consumption.** The current World Health Organization guidelines recommend limiting caffeine intake to less than 300 mg per day during pregnancy (Guilbert, 2003). However, there is a lack of clear evidence linking coffee consumption to adverse pregnancy outcomes. Multiple meta-analyses and cohort studies previously reported causal associations between maternal caffeine intake and adverse pregnancy outcomes, mainly focusing on spontaneous miscarriages (Fernandes et al., 1998; Greenwood et al., 2014; Li et al., 2015; Weng et al., 2008) and stillbirth (Bech et al., 2006; Gaskins et al., 2018; Greenwood et al., 2014; Matijasevich et al., 2006; Wisborg et al., 2003). However, the recent analysis using MR approach, of coffee consumption and pregnancy outcomes did not show significant causal relationship (**Table 3**) (Brito Nunes et al., 2022). The causal effect of caffeine consumption on miscarriage outcome was shown in a trans-ancestral GWAS meta-analysis of sporadic and recurrent miscarriages within the 23andMe dataset (Reynoso et al., 2024). This study used MR analysis with 127 SNPs associated with caffeine consumption as instrumental variables (**Table 3**) (Reynoso et al., 2024). Conversely, in MR study examining maternal coffee consumption and its effects on miscarriage, stillbirth, or pre-term birth, conducted through a GWAS meta-analysis by the Coffee and Caffeine Genetics Consortium, there was no impact of maternal coffee consumption on the risk of sporadic miscarriage (**Table 3**) (Brito Nunes et al., 2022). Overall, the evidence remains inconclusive, indicating the need for further research to clarify the relationship between caffeine consumption and miscarriage.

**Alcohol consumption.** Higher alcohol consumption was widely reported as related to negative pregnancy outcomes, and numerous studies reported associations between alcohol intake and miscarriage risk (Chai et al., 2022; Sundermann et al., 2019; Xu et al., 2023). However, together with many observational studies that showed association between alcohol consumption and miscarriage risk, there are also studies that cannot draw a direct association between alcohol intake and increased risk of miscarriages (Ng et al., 2021; Skogsdal et al., 2022). Observational studies are not consistent in their results due to the challenges with collecting and quantifying the alcohol intake, since there is no consensus on the classification of types of alcohol and the number of drinks consumed of every kind of alcohol. MR studies performed to evaluate the causal effects of alcohol consumption and the risk of sporadic or recurrent miscarriages demonstrated no association between the genetic variants for alcohol intake and miscarriage events (**Table 3**) (Laisk et al., 2020; Painter et al., 2021; Yuan et al., 2021). A recent study examining the associations of smoking, alcohol, and coffee consumption with PL reported no significant link between genetically predicted moderate alcohol intake and the risk of PL, which includes stillbirth, spontaneous miscarriage, and pregnancy termination (Yuan et al., 2021). The odds ratio (OR) for alcohol consumption was 1.09 (95% CI: 0.93–1.27), suggesting no statistically significant effect of moderate alcohol intake on PL. However, as this study did not distinguish between types of PL, further research is needed to assess genetic associations specifically between alcohol consumption and stillbirth. Such results from both observational studies and MR analyses indicate a need for additional larger exploration exploring effects of alcohol consumption on PL outcomes.

**Maternal age** is an important factor associated with an higher risk of miscarriage (Carroli et al., 2001). Several studies demonstrated a compelling association between maternal age and the risk of miscarriages (Andersen et al., 2000; George et al., 2006; Khalil et al., 2013; Magnus et al., 2019; Wilcox et al., 1988). For example, some of these studies found a progressive increase in miscarriage rates with advancing maternal age, particularly beyond 45 years old (Andersen et al., 2000; Magnus et al., 2019). Furthermore, a population-based case-control study published highlighted a nonlinear relationship, indicating that both very young and older maternal ages are linked to heightened miscarriage risks (George et al., 2006). The same pattern was observed between the maternal age and the occurrence of stillbirth (Flenady et al., 2011; Huang et al., 2008; Reddy et al., 2006). Several studies describe the relationship between maternal age and stillbirth risk as U-shaped, with increased risks at both extremes of maternal age (Gardosi et al., 2013; MacDorman & Gregory). While no MR study currently assessed the causal relationship between the sequalae of women’s reproductive factors, existing **epidemiological evidence** underscores the critical influence of maternal age on miscarriage outcomes, informing age-specific healthcare interventions. Future MR studies would help to shed the light on causal associations between the observed trend in stillbirth cases.

**Family history of miscarriage.** Large-scale cohort epidemiological studies research highlight the familial aggregation of miscarriage risk (Andersen et al., 2000; George et al., 2006; Khalil et al., 2013; Magnus et al., 2019; Wilcox et al., 1988; Woolner & Bhattacharya, 2023). A family history of miscarriages is strongly associated with an increased risk of PL in epidemiological studies, suggesting a genetic predisposition. Further research in this area could provide more insights into the genetic components underlying miscarriage risks, particularly by integrating family history data into genetic studies using MR methodology.

**Psychological stress.** The epidemiological relationship between psychological stress or depression and miscarriage has been extensively studied. Overall, prior psychological stress and depression may increase miscarriage risk, possibly through physiological mechanisms such as elevated cortisol levels (Almeida et al., 2016; Qu et al., 2017). Yunan He et al. identified a causal relationship between four mental disorders—anxiety, major depressive disorder, bipolar disorder, and insomnia— and spontaneous abortion using MR analyses. However, no causal link was found between broad depression and spontaneous abortion. Additionally, they found no evidence of a causal relationship between these five common mental disorders and recurrent miscarriage (He et al., 2024). No causal relationship between depression, dysthymia, and spontaneous abortion has been established in similar MR study (Ling et al., 2024) (**Table 3**).

## Discussion

Recent large-scale studies revealed 30 variants reaching genome-wide significance marking an important step forward in understanding the biology of pregnancy loss and its outcomes. New GWAS associations with PL outcomes accentuate the complex nature of pregnancy and miscarriage mechanistic pathophysiology. Functional investigation of mechanisms by which the genes in associated PL loci operate could pave the way for targeted therapies and diagnostic approaches aimed at addressing reproductive loss. Such understanding also holds promise for advancing precision medicine strategies tailored to individuals at risk of miscarriage. Such functional studies evaluating GWAS results should be performed to ensure the credibility of statistical associations. Additional studies and replication in wider number of ancestries is required, to account for diversity between populations due to history, geographical influences, population size bottlenecks and genetic drift (Beaty et al., 2005; Kraft et al., 2009; Lappalainen et al., 2024; Sawyer et al., 2005). Our investigation into genetic relationships between higher pregnancy loss PGS and diaphragmatic hernia, eosinophilic esophagitis, and asthma with exacerbation outcomes through PheWAS for PGS bring novel highlights into PL pathophysiology and its risk factors warranting further investigation.

The investigation into the causality between a range of external risk factors and miscarriage, stillbirth and pregnancy loss generally are a key important point to push further previous epidemiological associations. Potential risk factors for pregnancy loss include caffeine and alcohol consumption, smoking, maternal age, psychological stress or depression, BMI, and family history of miscarriages. Our ability to use MR prove or disregard causality will fill in significant gaps in current research. Observational studies often indicate correlations between these traits and adverse pregnancy outcomes. However, MR studies with larger PL GWAS will provide a more robust framework for identifying potential causal relationships by minimising confounding factors.

Caffeine consumption, higher BMI, and smoking show some support for causal effects of miscarriage risk, particularly in studies employing extensive genetic data from diverse populations. However, the evidence is less clear for traits such as alcohol consumption, maternal age, psychological stress, and family history of miscarriage, where MR studies either do not demonstrate causality or are limited in scope. The association between a family history of miscarriages remains underexplored in MR studies, despite epidemiological evidence suggesting genetic predisposition.

The complexity of genetic influences on pregnancy outcomes necessitates further research. Future studies should aim to expand the genetic data available for these traits and improve the statistical power of MR analyses. Additionally, integrating diverse populations and considering gene-environment interactions will be crucial in unravelling the genetic architecture underlying pregnancy loss outcomes. Continued exploration in this domain holds the potential to inform more effective preventative strategies and personalised medical interventions for women at risk.

## Author contributions

Z.M. Designed and led the manuscript write-up and literature review. Proofread and prepared submission ready manuscript and submission package. VP. Led in literature review and manuscript writing, Designed and conducted Polygenic Score (PGS) and Phenome-Wide Association Study (PheWAS) analyses, including comprehensive data interpretation. S.A., U.Y., K.R. Led in literature review and manuscript writing. Ch.Chu., Ch.S., A.R., Y.K., S.N., M.N., G.A., M.S., L.K., G.E., H.M., M.R. Contributed to literature review and writing the manuscript. F.N. Managed, and coordinated the project. Abr.A. Designed, managed, and coordinated the project. Contributed to the literature review and manuscript writing. I.P. Co-principal investigator of the project. Designed and supervised the project and the manuscript write-up, literature review. Proofread and edited submission ready manuscript. As guarantor, had full access to all study data and was responsible for its integrity and analysis accuracy. D.D., Sh.T. Designed, managed, and coordinated the project. Y.Sh. Designed and supervised the manuscript write-up. Designed and conducted Polygenic Score (PGS) and Phenome-Wide Association Study (PheWAS) analyses, including comprehensive data interpretation. Proofread submission ready manuscript. A.A. Principal investigator of the project. Designed and supervised the manuscript write-up. As guarantor, had full access to all study data and was responsible for its integrity and analysis accuracy.

## Funding

This publication has been produced within the framework of the Grant Advanced pregnancy loss study in Uzbekistan – “ALSU” (REP-03032022/192), funded under the MUNIS Project, supported by the World Bank and fundamental grant PZ-20200930489, sponsored by the Government of the Republic of Uzbekistan. The statements do not necessarily reflect the official position of the World Bank and the Government of the Republic of Uzbekistan. The research of YSh was funded by EMBO SLG 5423-2023.

The analyses within UK Biobank were conducted under approved project n.75072; project PI-YSh.

## Tables

**Supplementary Table 1.** PheWAS results between PL PGS and 280 ICD-10-defined disease outcomes in the Uk Biobank, including case/control counts, association model and association test statistic effect estimates.

## Supporting information

Supplementary Table 1. PheWAS results between PL PGS and 280 ICD-10-defined disease outcomes in the Uk Biobank

## Data Availability

All data generated in this study can be accessed by contacting the authors upon reasonable request.

